# Estimating Active Cases of COVID-19

**DOI:** 10.1101/2021.12.09.21267355

**Authors:** Javier Álvarez, Carlos Baquero, Elisa Cabana, Jaya Prakash Champati, Antonio Fernández Anta, Davide Frey, Augusto Garcia-Agundez, Chryssis Georgiou, Mathieu Goessens, Harold Hernández, Rosa Lillo, Raquel Menezes, Raúl Moreno, Nicolas Nicolaou, Oluwasegun Ojo, Antonio Ortega, Estrella Rausell, Jesús Rufino, Efstathios Stavrakis, Govind Jeevan, Christin Glorioso

**Affiliations:** IMDEA Networks Institute, Spain; U. Porto & INESC TEC, Portugal; Univ Rennes, IRISA, CNRS, Inria, 35042 Rennes, France; Brown U., USA; U. of Cyprus, Cyprus; Consulting, France; U. Carlos III de Madrid, Spain; U. Minho, Portugal; Madox Viajes, Spain; Algolysis Ltd., Cyprus; IMDEA Networks Institute & U. Carlos III de Madrid, Spain; U. Southern California, USA; U. Autónoma de Madrid, Spain; Academics for the Future of Science, Inc. & DICE Institute, Pathcheck Foundation, USA; Academics for the Future of Science, Inc., University of California San Francisco, & DICE Institute, Pathcheck Foundation, USA

## Abstract

Having accurate and timely data on active COVID-19 cases is challenging, since it depends on the availability of an appropriate infrastructure to perform tests and aggregate their results. In this paper, we consider a case to be active if it is infectious, and we propose methods to estimate the number of active infectious cases of COVID-19 from the official data (of confirmed cases and fatalities) and from public survey data. We show that the latter is a viable option in countries with reduced testing capacity or infrastructures.

## 1 INTRODUCTION

The COVID-19 disease epidemic started having a deep impact in societies across the world as the virus spread globally in early 2020. During its initial months, the knowledge about the virus’s properties and about the symptoms of COVID-19, and the ability to test for the disease were limited [23]. These factors made obtaining accurate estimates of cases challenging. This problem affected the estimation of a number of important metrics, including the number of daily, cumulative, and active infectious cases. The limitations of these estimations motivated alternative approaches such as estimating cases based on fatalities [32], direct surveys [10], and indirect reporting [25].

In this article, we compare several approaches to estimating the number of active cases in different countries. We consider a case to be *active* if it is *infectious*. We include methods that use and correct official reporting data of COVID-19 cases and deaths [20, 34] as well as survey-based approaches. The latter may rely on direct survey questions that can be used to determine if the participant is an active symptomatic case of COVID-19. Alternatively, they can use questions in which participants indirectly report the number of cases among their contacts, and use the Network Scale-up Method [3, 13] to obtain the estimate. In this work, we use data from the COVID-19 Trends and Impact Surveys (CTIS) deployed by the U. of Maryland and Carnegie Mellon University with the support of Facebook [1, 10]. Indirect reporting data is also extracted from the responses collected by the CoronaSurveys project [2, 7]. We observe how different methods correlate in some countries while they greatly differ in others, and explore the possible reasons for these discrepancies.

## 2 METHODS

### 2.1 Official Reporting: Confirmed Active

For official data, we refer to daily reported cases based on PCR testing, collected from data aggregates such as those provided by Johns Hopkins University [20] or University of Oxford [4]. From this data, an official number of active cases could be computed by subtracting the number of fatalities and recovered subjects from the number of confirmed cases [34]. Unfortunately, the number of recovered cases from these sources is unreliable or unavailable in some countries (e.g., Greece, Spain).

In order to have an estimation of the number of active cases from the official numbers, we use the following approach: we assume that each reported case is active for a fixed period, namely the number of days a case is infectious. To estimate the usual active duration of a COVID-19 case, we assume a case is infectious for 3 days during the incubation period (until symptom onset) and for 7 days after symptom onset [6, 16, 17, 29]. This yields that a case is considered active for *α* = 10 days. This parameter is also applied to asymptomatic cases, since they show similar RT-PCR cycle thresholds^1^ (Ct) [29]. Then, we obtain the estimate of active cases from confirmed cases, which we label as *Confirmed Active*, by extending each confirmed case over *α* days.

### 2.2 Fatality-based estimation: CCFR and CCFR Fatalities

On March 13 2020, a news article in Nature [23] brought attention to the high level of under-detection of COVID-19 and suggested the use of estimates of the case fatality rate (CFR) to infer the likely number of cases. Also in March 2020, the Centre for the Mathematical Modelling of Infectious Diseases at the London School of Hygiene & Tropical Medicine made available a more detailed estimation of the level of under-detection of COVID-19 symptomatic cases [27, 28]. This study took into account the delay distribution from symptoms to death, and used a more precise estimate of the CFR from an ongoing study with Wuhan data that established it around 1.38% [32]. In this article, we use this CFR even though it represents a simplification, since the CFR will vary geographically based on government tracking and medical capabilities, as well as country demographics.

Following the technique in [28], it is possible to estimate the actual cumulative number of symptomatic cases at time *t* by looking at the reported cumulative number of total cases and deaths, and calculating the apparent CFR_*t*_. If this CFR_*t*_ is higher than the baseline CFR_*b*_ = 1.38%, assuming it is accurate, this is an indication of under-detection of cases. Conversely, the actual number of cumulative cases can be obtained by multiplying the cumulative reported cases by 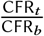. In [24], we provided a detailed description of this estimation and showed that it matched the serology-based prevalence observed in Spain [14].

Let us now consider the relationship between the daily and the cumulative number of cases. Let *D* =(*d*_1_, …, *d*_*n*_) be the series of daily new cases and *C* = (*c*_1_, …, *c*_*n*_) the respective cumulative number of cases, for *n* consecutive days since the start of the pandemic. Then, *d*_*i*_ = *c*_*i*_ − *c*_*i*− 1_ with *c*_0_ = 0. We observe that, if *C* is not strictly non-decreasing, the *D* series may exhibit negative values (it is a rare event, but some countries have resorted to negative values in daily cases and/or deaths to correct past misreporting).

As we mentioned above, if the *D* and *C* series are based on the confirmed data, they can underestimate the true number of cases [23]. The CFR-based techniques that we developed in [24] provide a series *Ĉ* that estimates the number of cumulative cases. However, this series is not always non-decreasing, and needs some processing in order to estimate the likely true number of daily cases. Our target is to derive the series of estimated daily cases 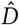. To this end, we consider 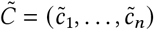 where 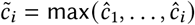, so that the cumulative series 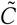 is always non-decreasing. Now, we can define 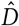 such that 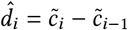 with 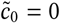.

As an additional reference, we also define a very simple estimator of daily cases 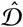 that only takes into account the daily number of reported deaths. Let *F* = (*f*_1_, …, *f*_*n*_) be the series of daily fatalities attributed to COVID-19. We define 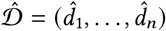 such that 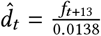, where *t* is a day of reference and *t* + 13 is a day 13 days later. That is, this considers the number of deaths in a given day and uses it to infer how many cases likely occurred 13 days before, by using as reference the Wuhan CFR_*b*_ = 1.38%. The delay of 13 days is the median time from onset of symptoms to death, estimated from the CDC COVID-19 Pandemic Planning Scenarios [12]. The disadvantage of this technique is that it cannot predict the daily cases in the most recent 13 days, while the advantage is that it only requires an accurate reporting of COVID-19 deaths, and does not need the series of detected cases.

From these estimates we derive two series of active cases: CCFR, derived from 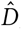, and CCFR Fatalities, derived from 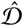. Both series are obtained by considering each daily case as active for the next *α* = 10 days, and letting it expire after that time, so that it no longer contributes to subsequent estimates of active cases. This transformation also helps smooth the data, since cumulative cases combine information from multiple days and reduce the effect of daily noise. Note that both estimates are of *symptomatic* active cases, since we use the CFR to obtain them.

### 2.3 Survey-based Direct Reporting: UMD CLI and UMD CLI WHO

Since the spring of 2020, Facebook has been collaborating in the context of their program Data for Good [10] with CMU and the U. of Maryland promoting surveys created by these two institutions [21]. Both surveys collect thousands of daily responses, in which participants report, among other elements: symptoms, habits, or vaccination status. The *Delphi Group at Carnegie Mellon University U*.*S. COVID-19 Trends and Impact Survey in partnership with Facebook* (Delphi US CTIS) [8] collects around 50,000 responses daily in the USA, while the *U. of Maryland Social Data Science Center Global COVID-19 Trends and Impact Survey in partnership with Facebook* (UMD Global CTIS) [11] collects more than 100,000 responses daily from many countries in the world. For this article, we have used the individual responses of UMD Global CTIS to estimate the total number of active cases using both direct and indirect survey questions.

We observed that the set of responses to the UMD Global CTIS has to be curated. As a first approach, we remove all responses that declare to have all 12 symptoms in the last 24 hours. We also remove responses with high values (more than 100) in certain questions.^2^ More specifically, we remove the survey responses that:

- Declare to have all 12 symptoms in the last 24 hours (fever, cough, difficulty breathing, fatigue, stuffy or runny nose, aches or muscle pain, sore throat, chest pain, nausea, loss of smell or taste, eye pain, headache).
- Declare at least one symptom and give a high value to the question “For how many days have you had at least one of these symptoms?”
- Answer *Yes* to the question “Do you personally know anyone in your local community who is sick with a fever and either a cough or difficulty breathing?” and give a high value to the question “How many people do you know with these symptoms?”
- Give a high value to the question “How many people slept in the place where you stayed last night (including yourself)?”
- Give a high value to the question “How many years of education have you completed?”

After filtering, each survey response is considered to be a *COVID-like-illness* active case if it declares the following combination of symptoms: fever, along with cough, shortness of breath, or difficulty breathing. Let *cli*_*t*_ be the number of *COVID-like-illness* active cases among the *k*_*t*_ survey responses on day *t*, we estimate the ratio of active cases on that day as *cli*_*t*_ /*k*_*t*_, which is labeled UMD CLI. Similarly, a response is considered to be an active case of *COVID-like illness according to the WHO*, if it declares the following symptoms: fever, cough, and fatigue [33]. If there are *cliWHO* such responses, the corresponding ratio is obtained as *cliWHO*_*t*_ /*k*_*t*_ and is labeled UMD CLI WHO. Both UMD CLI and UMD CLI WHO are estimates of the ratio of active symptomatic cases in the population.

### 2.4 Survey-based Indirect Reporting: Coronasurveys and UMD CLI Local

In [2, 13], we present CoronaSurveys, a survey-based estimate using indirect reporting and the Network Scale-up Method (NSUM) [3, 22]. In the CoronaSurveys poll, we have the following questions relative to a pre-selected geographical area: (1) “How many people do you know personally in this geographical area?” (2) “How many of the above have been diagnosed or have had symptoms compatible with COVID-19, to the best of your knowledge?”. The first question provides the *reach, r*_*i*_, variable of a participant, *i*, and the second the number of *cases, c*_*i*_. The ratio ∑_*i*_ *c*_*i*_ / ∑_*i*_ *r*_*i*_ provides an estimator of the fraction of the population that has had COVID-19. In order to estimate the number of active cases, we included the question (3) “How many are still sick?”. If the answer by participant *i* is *s*_*i*_, an estimator of the fraction of the population that currently has COVID-19 is ∑_*i*_ *s*_*i*_/ ∑_*i*_ *r*_*i*_. We label this estimate of the ratio of active cases CoronaSurveys. The UMD Global CTIS also implements indirect reporting via two questions:

- B3: “Do you personally know anyone in your local community who is sick with a fever and either a cough or difficulty breathing?”
- B4: “How many people do you know with these symptoms?”

The combination of both allows us to extract the number of cases of *COVID-like illness* in the local community of a participant *i*, denoted *ccli*_*i*_. This information is similar to the number of sick cases *s*_*i*_ provided by the third question above in the CoronaSurveys poll.

Unfortunately, the UMD Global CTIS does not include a direct question that collects from participants the size *r*_*i*_ of their local community. Instead, we estimate the average size of the local community, 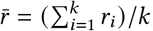, from the *k* = ∑_*t*_*k*_*t*_ available survey responses in the data set. To this end, we first compute the direct ratio *cli*/*k* of *COVID-like-illness* active cases in the *k* responses as described above (UMD CLI), where *cli* = ∑_*t*_ *cli*_*t*_. Then, we assume that the direct ratio *cli*/*k* of active cases must be equal to the ratio 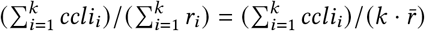 obtained using the indirect reporting. Hence, we estimate 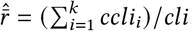. Finally, we estimate the ratio of the population that is sick with a COVID-like illness on day *t* from the *k*_*t*_ responses to the UMD Global CTIS on that day as 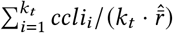. We label this estimate UMD CLI local.

## 3 RESULTS

To compare the discussed methods, we present our estimates computed for Greece, the UK, Brazil, India, Peru and Ecuador from January 1st to May 15th of 2021 in Figures 1a to 2c. The plot for Brazil shows the raw values as obtained with the methods described above, and the results of smoothing these values with a Gaussian kernel. The rest of the plots show only the smoothed curves for clarity. The set of countries was selected to have a combination of countries in which the estimates are close to each other (Greece and the UK), and countries in which there is large disparity from different geographical areas (Southamerica and Asia)^3^. We exclude the CoronaSurveys method from these figures because the number of survey responses in these countries is too low in the considered period. We generally observe that the estimated numbers of active cases using the confirmed (reported) data are lower than those derived from surveys. As a comparison for indirect reporting with the CoronaSurveys method, Figure 3 presents the estimates of active cases for the region of Madrid in Spain. The number of survey responses used for the estimates and their reach is presented in Table 1. A comparison of the ratio of cases of some of the methods with the excess mortality and the test positive rate (TPR) is provided in Table 2.

**Table 1.**
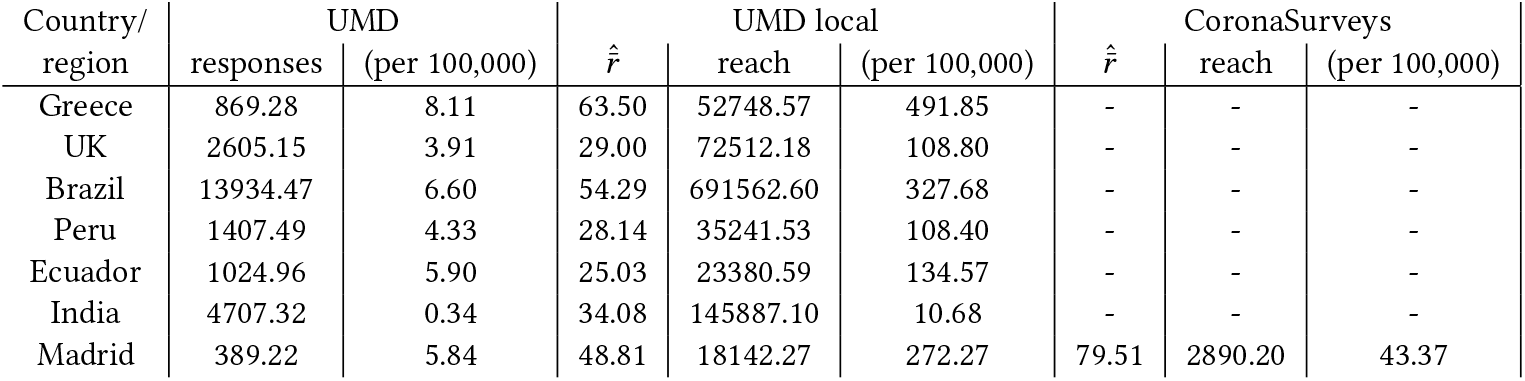
Average daily sample size (direct or indirect) used by each of the survey-based estimates, and the fraction of the population represented (in persons per 100, 000). *UMD responses* are the average daily number of responses used for the direct estimates *UMD CLI* and *UMD CLI WHO. UMD local reach* is the estimated average daily reach used for the indirect estimate *UMD CLI local. CoronaSurveys reach* is the estimated average weekly reach used for the indirect estimate *CoronaSurveys*.

**Table 2.** Ratio of different estimates (namely, average value of *CCFR, UMD CLI, UMD CLI local, CoronaSurveys*) with respect to the average value of *Confirmed active* (period Jan 1st to May 15th, 2021), compared with (1) the ratio of excess mortality, from The Economist [31] and IHME [18], with respect to official COVID mortality (period March 2020 to May 2021), and (2) the test positive rate (TPR) from Our World in Data [15] (OWID) and the UMD Global CTIS [21] (period Jan 1st to May 15th, 2021). For Madrid, official the official excess mortality is obtained from [19] (period March 2020 to May 2021) and the official TPR from [5] (period Jan 1st to May 13th, 2021).

| Country/<br>region | Confirmed<br>active<br>(per 100,000) | CCFR<br>ratio | UMD<br>CLI<br>ratio | UMD<br>CLI local<br>ratio | Corona<br>Surveys<br>ratio | Excess mortality<br>ratio |  | Test positive<br>rate (TPR) |  |
| --- | --- | --- | --- | --- | --- | --- | --- | --- | --- |
|  |  |  |  |  |  | Economist | IHME | OWID | UMD |
| UK | 219.84 | 1.64 | 4.12 | 3.72 | - | - | 1.0 to 1.1 | 2.65% | 6.13% |
| Greece | 163.68 | 2.14 | 3.33 | 3.32 | - | 0.49 | 1.25 to 1.5 | 4.13% | 4.38% |
| Brazil | 277.64 | 2.23 | 6.63 | 6.55 | - | 1.06 | 1.46 | - | 44.51% |
| Peru | 196.84 | 7.35 | 23.16 | 23.23 | - | 2.66 | 2.36 | 16.73% | 27.68% |
| India | 78.05 | 0.79 | 23.94 | 21.55 | - | - | 2.97 | 7.10% | 22.71% |
| Ecuador | 83.99 | 2.16 | 45.30 | 45.15 | - | 2.95 | 2.5 to 3 | 31.86% | 26.49% |
| Madrid | 328.86 | 0.86 | 2.19 | 1.91 | 2.76 | 1.40 [19] |  | 11.32% [5] | 13.63% |

**Fig. 1.**
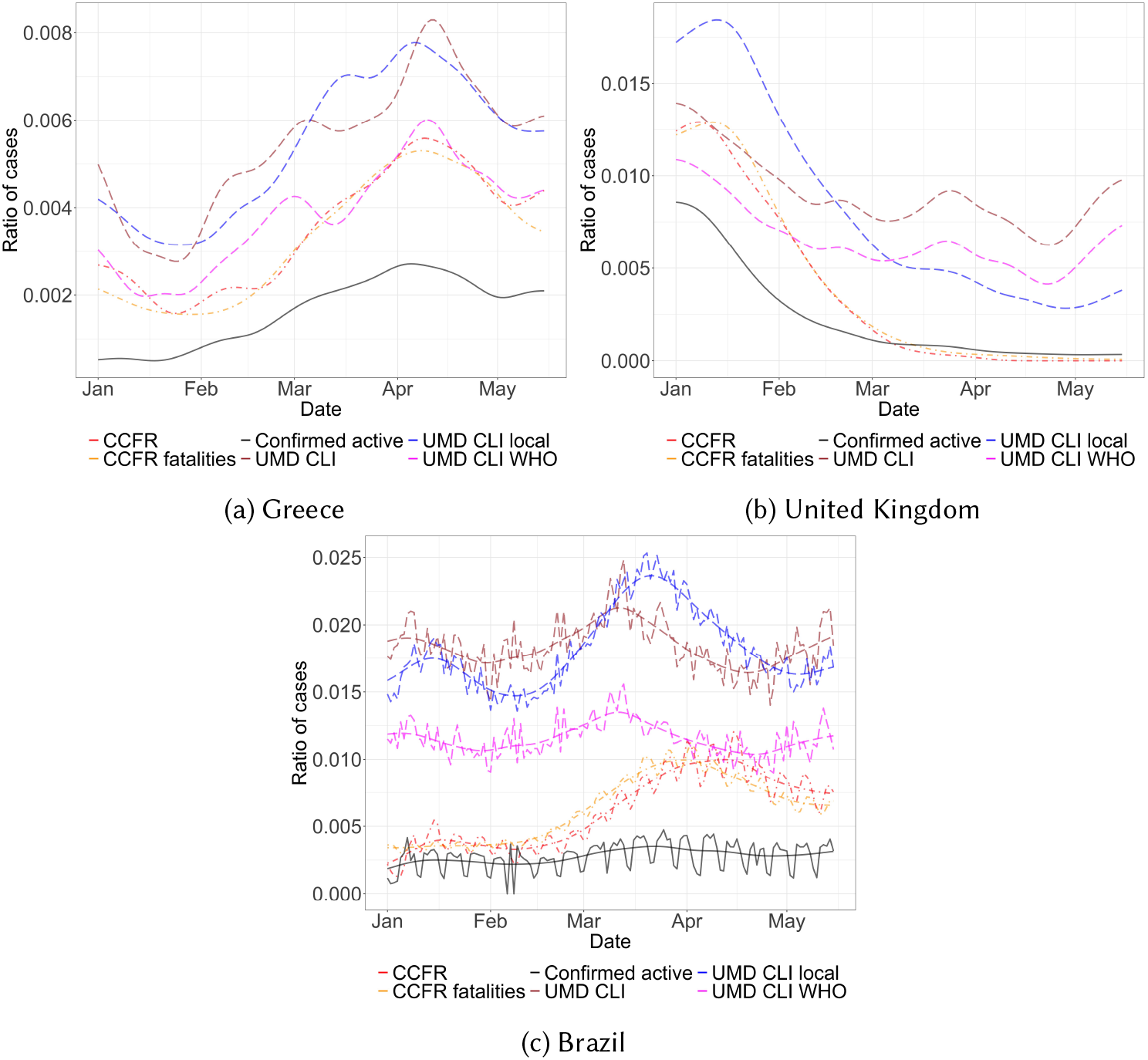
Active cases in Greece, the UK, and Brazil in 2021. In the plot for Brazil the raw daily estimates are shown together with curves resulting of applying Gaussian smoothing (bandwidth 15). In the other plots all curves have been smoothed.

## 4 DISCUSSION

For Greece (Figure 1a), all estimates have the same behavior over time, but with different magnitudes. The estimate based on the confirmed cases, *Confirmed active*, appears clearly lower than the rest. Its values are roughly half of those obtained from fatalities (i.e., *CCFR* and *CCFR fatalities*). This difference may be due to better data on the number of fatalities than on the number of cases^4^. Similar trends, although with slightly larger values, can be observed for the estimates obtained from the survey responses *UMD CLI, UMD CLI WHO*, and *UMD CLI local*. Again, this difference can be due to under-reporting of fatalities, or to the fact that the symptomatic active cases identified with the surveys are over-counting the number of active infectious cases.

In the UK (Figure 1c), the estimates obtained from fatalities (*CCFR* and *CCFR fatalities*) follow each other very closely. Moreover they remain higher than those based on the confirmed cases *Confirmed active* between January and March, even if the difference is less dramatic than in Greece (the average ratio between *CCFR* and *Confirmed active* is of 1.64 in the UK and 2.15 in Greece, see Table 2). After March 2021, the number of cases dropped (partly thanks to vaccination) and fatality-based estimates tend to follows the confirmed-active one. Like for Greece, survey-based estimates reach higher values, but interestingly they remain high even when the other estimates are close to 0 (between March and May). Since the virus appeared relatively under control in that period in the UK, these high values probably results from the similarity between the symptoms of COVID-19 and those of other diseases. Interestingly the *UMD-CLI-local* estimate reaches higher values than the other survey estimates before March and lower values after March. This could be due to the fact that in periods of high epidemic activity people tend to share more information about their symptoms with friends and non-immediate family, and this is captured by the Network Scale-up method.

Brazil (Figure 1c) exhibits a different trend, where the estimates based on fatalities have a different shape and are much larger than the estimates based on confirmed cases. Such behavior suggests that fatalities are reported much better than confirmed cases. On the other hand, estimates based on surveys suggest a high under-reporting of confirmed cases, resulting in almost ten times more estimated active cases than officially confirmed cases. The *UMD-CLI-local* estimate appears to vary in a similar manner to the fatality-based estimates, but it appears to peak earlier, possibly suggesting delays in the reporting of deaths and confirmed cases. The other survey estimates peak even earlier but exhibit less prominent variations.

For India (Figure 2a), the curves obtained from confirmed cases and from fatalities appear to align, suggesting that the fatality ratio in the country matches the ratio of the baseline percentage from Wuhan. The estimates coming from the surveys however match the trend we observe in Brazil, suggesting that active cases are at least one order of magnitude higher than those estimated by confirmed cases. This possibly indicates significant under-reporting in confirmed cases and fatalities.

**Fig. 2.**
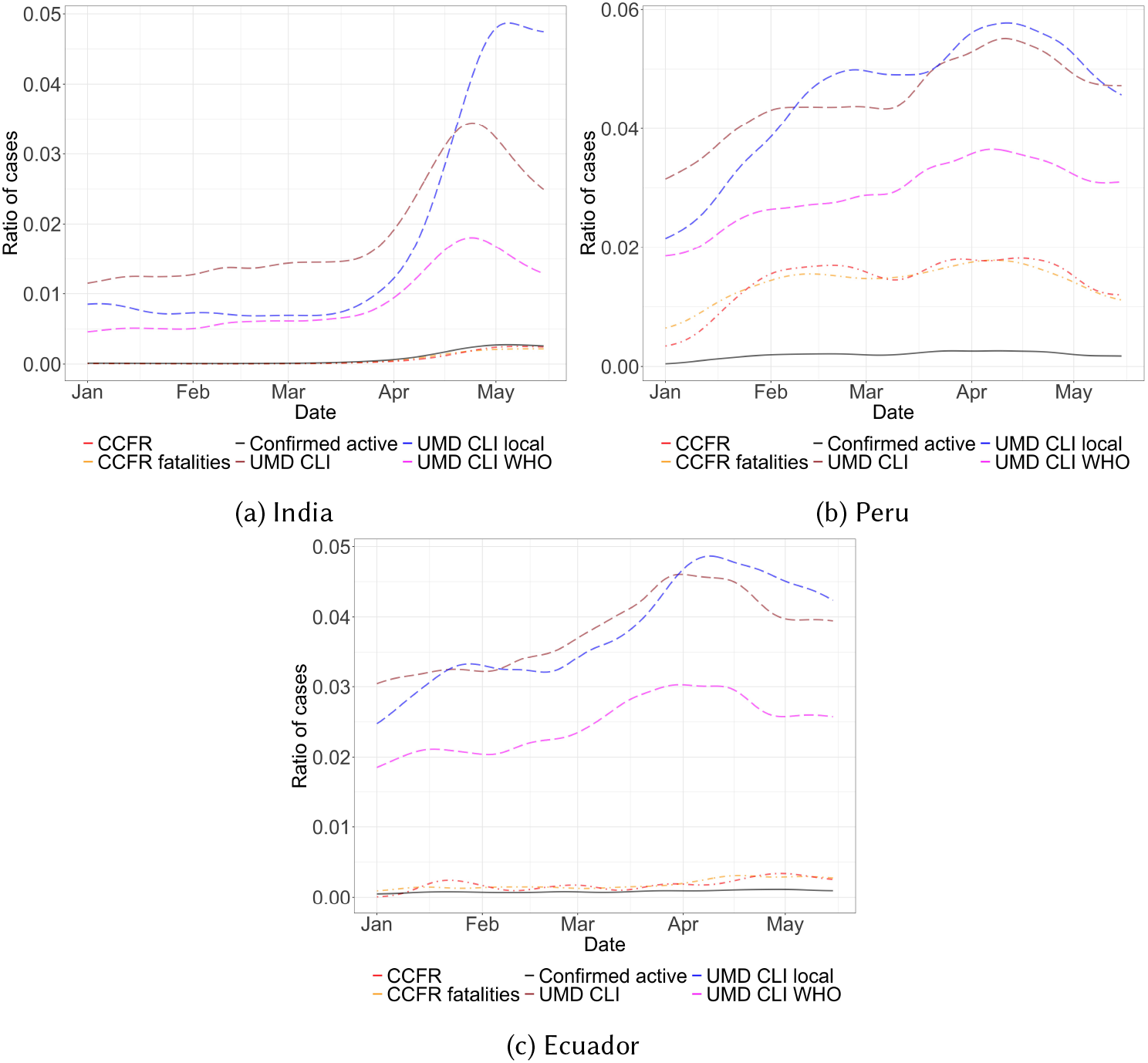
Active cases in India, Peru, and Ecuador in 2021. The curves are obtained by Gaussian smoothing.

Peru (Figure 2b) shows a significant difference between confirmed cases and fatality-based estimates. The other surveys-based estimates indicate a number of cases twice or three times as high as the fatality-based values, also indicating a significant degree of under-reporting.

Ecuador (Figure 2c) shows the largest discrepancy between official-data-based estimations and surveys. As we discuss further below, we believe the high excess mortality indicates a very significant degree of under-reporting in official data. As we observed in other countries, all survey-based estimates peak earlier than those based on official data. *UMD CLI* and *UMD CLI WHO* peak the earliest while *UMD CLI local* exhibits higher variations.

Regarding Figure 3, the estimates follow similar patterns, correctly identifying the burst of cases that Madrid suffered in January of 2021. It is worth mentioning that CoronaSurveys uses an average of 36 responses per day while the other survey-based estimates have close to 500 responses per day. Table 1 summarizes the daily number of responses, population coverage, and reach used for the different survey-based methods. As can be seen, the UMD survey receives at least 4 responses per 100,000 people on a daily basis in all the considered countries except India (possibly due to its large population and lower Facebook penetration). This is a good sample to get reliable estimates. Now, when indirect reporting is used, as can be observed the coverage of the population is much larger (up to 4.9 in a 1000 in Greece).

**Fig. 3.**
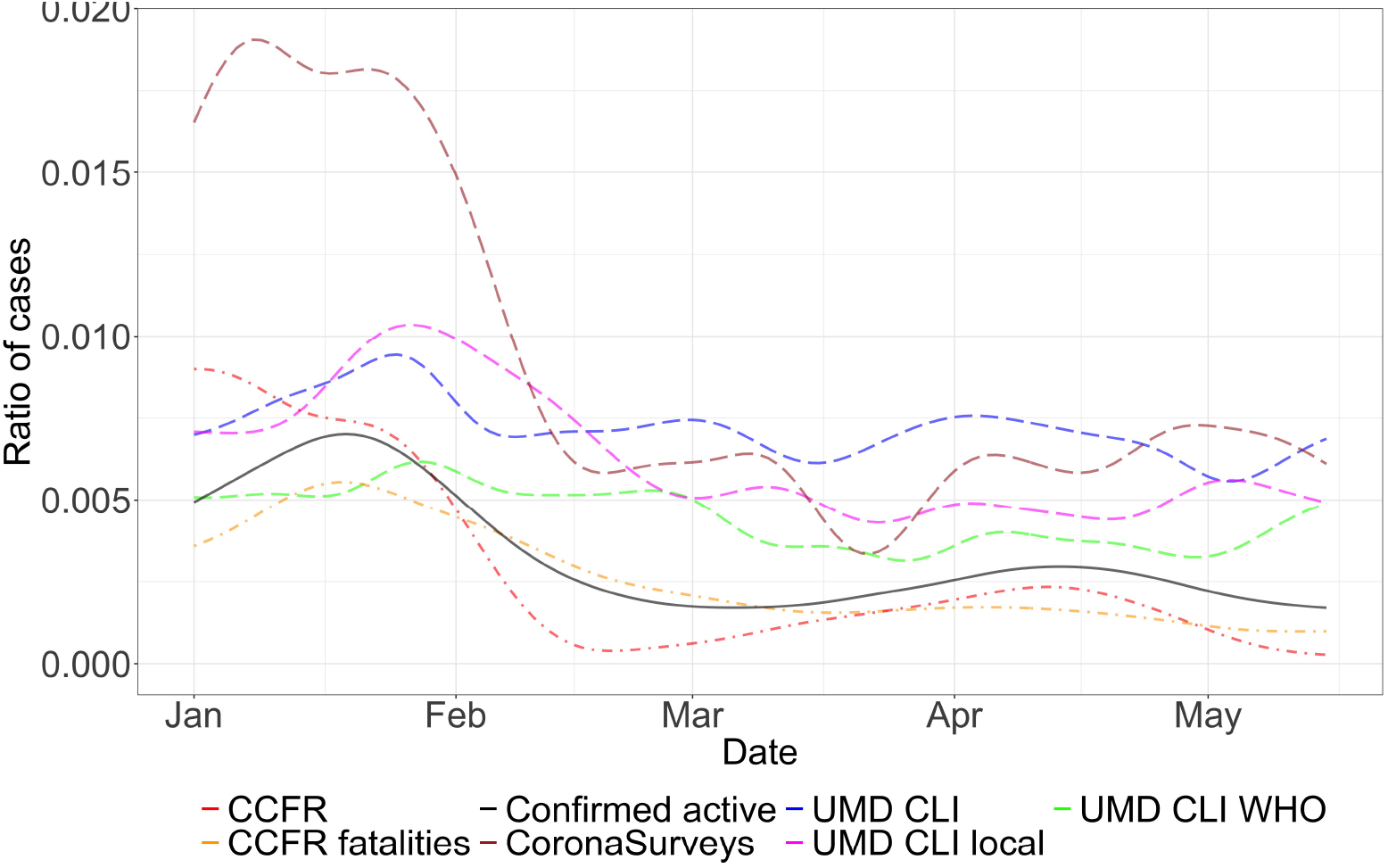
Active cases in Madrid, Spain, in 2021. The curves are obtained by Gaussian smoothing.

Finally, Table 2 compares several average daily metrics for the period Jan 1st to May 15th 2021. Column *Confirmed active* shows the average daily number of active cases estimated from the official number of new cases. *CCFR ratio* shows the ratio between the *CCFR* and *Confirmed active* estimates. Since the *CCFR* estimate relies on reported fatalities, a ratio larger than one suggests that fatalities may be better reported than cases. This is the case in Greece, Brazil, and Ecuador with values just above two, and especially Peru which has a much larger ratio. *UMD CLI ratio, UMD CLI local ratio* and *CoronaSurveys ratio* are the ratios of the corresponding average daily estimates with respect to *Confirmed active*. These ratios reflect whether the survey-based estimates are close to the official data. These ratios are moderately low for the UK, Madrid, and Greece, and slightly higher for Brazil. They are, in contrast, large in Peru, India, and Ecuador, hinting that official data in these countries may not be accurate. This seems to be confirmed by the fact that these countries show a higher ratio between the estimated *Excess mortality* and the official data on fatalities (shown in two columns with data from two different sources). A final metric shown in Table 2 is the *Test positive rate* (TPR), obtained from official sources [5, 15] (*OWID* column) and from direct responses to the UMD survey (*UMD* column). While the TPR from the official data is not very consistent with the previous observations, the TPR from the UMD survey is clearly high for Peru, India and Ecuador. Somewhat puzzling is the high value of roughly 45% for the TPR in Brazil, a country that has been heavily affected by the pandemic but that did not exhibit such high values for the previous ratios.

## 5 CONCLUSIONS AND FUTURE WORK

Symptom survey data appears to be a useful surrogate measure for active infectious COVID-19 cases, especially in countries where direct testing is not adequate or reliable. Disseminating surveys is also a relatively cost-effective intervention that does not rely on implementation by host countries. Having reliable estimates of true active infectious cases is important for predictive modeling and policy planning.

There are a number of limitations to our analyses and estimates. The techniques developed in Section 2.2 take as a reference the baseline CFR_*b*_ = 1.38% from Wuhan. This baseline assumed a population where susceptibility was uniformly spread across age groups. Countries that have vaccination in progress typically vaccinate older age groups first, meaning the baseline case fatality ratio will decrease, since younger groups have less morbidity. Other factors such as comorbidities, virus variants, and the limitations of healthcare infrastructure may have also affected CFR values [30]. We plan to adjust for this factor in the future by considering CFR geographical variability taking into account country, age [26], vaccination, and virus variants.

Surveys based on symptoms have the drawback of being unable to detect asymptomatic cases. Additionally, as discussed on [13], an insufficient number of responses does not allow for reasonable estimates. If responses are significantly outdated, and thus not reactive to rapid changes in cases, they will also significantly underestimate active cases. Spurious responses are also a risk. We are exploring how to curate responses more effectively and how to have better average reach estimates. Additionally, the known biases of NSUM [22] need to be taken into consideration when using indirect reporting.

Finally, we are developing a data-driven approach to process UMD Global CTIS data to determine whether a response is an active case of COVID-19 from the set of symptoms it declares and its connection to test positivity (given that a subset of the respondents also reported the outcome of a diagnosis test). Here the aim is to search for a better combination of signals than those currently provided by UMD CLI and UMD CLI WHO.

The COVID-19 disease epidemic coincided with the availability of technology for the collection of worldwide symptom’s survey data across an extended period of time. With this study we have shown the possibilities brought forward by this new collection of data, and how it can be used to complement and cross-check more traditional mechanisms for health surveillance. We believe that the full potential of these data driven approaches is still untapped and can be applied to other targets of interest.

## Data Availability

All the data is available in the GitHub repository of the CoronaSurveys project https://github.com/GCGImdea/coronasurveys.

https://www.coronasurveys.org

https://github.com/GCGImdea/coronasurveys

## 6 ACKNOWLEDGEMENTS

We want to thank the whole CoronaSurveys Team [7] for the collective effort. We also want to thank Tamer Farag and Kris Barkume for useful discussions on the survey data. This work is partially supported by grant CoronaSurveys-CM, funded by IMDEA Networks and Comunidad de Madrid.

## Footnotes

1 Cycle threshold is the measure of the concentration of viral genetic material found in a sample using RT-PCR [9].

2 A threshold of 100 is conservative and filters out unrealistic responses, like those reporting millions.

3 Estimates for most countries are available at https://github.com/GCGImdea/coronasurveys.

4 However, it could also be due to an artifact of the methods described in Section 2.2, like the value CFR_*b*_ used.

